# Bipolar disorder and cannabis use: a bidirectional two-sample Mendelian randomization study

**DOI:** 10.1101/2020.06.11.20128470

**Authors:** Oskar Hougaard Jefsen, Maria Speed, Doug Speed, Søren Dinesen Østergaard

## Abstract

**Aims:** Cannabis use is associated with a number of psychiatric disorders, however the causal nature of these associations has been difficult to establish. Mendelian randomization (MR) offers a way to infer causality between exposures with known genetic predictors (genome-wide significant single nucleotide polymorphisms (SNPs)) and outcomes of interest. MR has previously been applied to investigate the relationship between lifetime cannabis use (having ever used cannabis) and schizophrenia, depression, and attention-deficit / hyperactivity disorder (ADHD), but not bipolar disorder, representing a gap in the literature.

**Methods:** We conducted a two-sample bidirectional MR study on the relationship between bipolar disorder and lifetime cannabis use. Genetic instruments (SNPs) were obtained from the summary statistics of recent large genome-wide association studies (GWAS). We conducted a two-sample bidirectional MR study on the relationship between bipolar disorder and lifetime cannabis use, using inverse-variance weighted regression, weighted median regression and Egger regression.

**Results:** Genetic liability to bipolar disorder was significantly associated with an increased risk of lifetime cannabis use: scaled log-odds ratio (standard deviation) = 0.0174 (0.039); *P*-value = 0.00001. Genetic liability to lifetime cannabis use showed no association with the risk of bipolar disorder: scaled log-odds ratio (standard deviation) = 0.168 (0.180); *P*-value = 0.351. The sensitivity analyses showed no evidence for pleiotropic effects.

**Conclusions:** The present study finds evidence for a causal effect of liability to bipolar disorder on the risk of using cannabis at least once. No evidence was found for a causal effect of liability to cannabis use on the risk of bipolar disorder. These findings add important new knowledge to the understanding of the complex relationship between cannabis use and psychiatric disorders.

## Introduction

Cannabis is the most widely used illicit drug worldwide (1). Cannabis produces acute psychiatric symptoms (2) and its use has been associated with the development of a range of psychiatric disorders, including schizophrenia (3, 4), depression (5), attention deficit hyperactivity disorder (ADHD) (6), and bipolar disorder (7). Understanding the relationship between cannabis use and the development of psychiatric disorders is important for effective prevention and intervention, however the causal nature of this relationship has been difficult to establish. Cannabis use might increase the risk of a psychiatric disorder, a psychiatric disorder (prodromal or manifest) might increase the risk of cannabis use, or the two might share common risk factors (confounding).

Determining the validity of these causal models is challenging, as observational data are sensitive to a multitude of confounders, and randomized controlled trials (RCTs) are unfeasible and unethical. Faced by this challenge, Mendelian Randomization (MR) – a method from genetic epidemiology – may be applied to infer causality (8). MR uses single nucleotide polymorphisms (SNPs) associated with an exposure of interest (e.g. cannabis use) as instrumental variables. Since SNPs are randomly assigned during meiosis, a significant association between the instrumental variables (SNPs) and the outcome (e.g. psychiatric disorder) supports a causal effect of the exposure of interest on the outcome. MR has previously been used to test the relationship between cannabis use and schizophrenia (9-11), depression (12), and ADHD (13), but not bipolar disorder, representing a gap in the literature. Patients with bipolar disorder show a high prevalence of cannabis use (14), however longitudinal studies on cannabis use and the risk of bipolar disorder have yielded mixed results (15-19). Both lifetime cannabis use (having ever used cannabis) and bipolar disorder are heritable traits with known genetic risk factors (SNPs)(9, 20), making bidirectional MR possible. We therefore aimed to increase the understanding of the causal relationship between bipolar disorder and cannabis use via MR, using results from recent large genome-wide association studies (GWAS).

## Methods

For the purpose of this study, we used summary statistics from two GWAS, namely that on lifetime cannabis use by Pasman et al. (9) and that on bipolar disorder by Stahl et al. (20). The study by Pasman used data from 184,765 individuals of European ancestry from the International Cannabis Consortium, 23andMe, and the UK-Biobank. Of these individuals, 28.8% had reported using cannabis at least once during their lifetime. The study by Stahl et al. used data from 198,882 individuals of European ancestry collected from 39 cohorts across Europe, North America and Australia. Of these individuals, 29,764 met DSM-IV or ICD-10 criteria for a lifetime diagnosis of bipolar disorder. Both studies provide summary statistics for the only most significant SNPs from their full GWAS, then genome-wide summary statistics from a smaller GWAS. In the study by Pasman et al., the smaller GWAS contained 162,082 individuals (of which 26.7% reported having ever used cannabis), while for Stahl et al., it contained 51,710 individuals (20,352 of whom had been diagnosed with bipolar disorder).

We investigated whether lifetime cannabis use is a causal risk factor for bipolar disorder. This required us to construct a genetic predictor for cannabis, containing SNPs for which we also had summary statistics for bipolar disorder. For this we used lifetime cannabis use-summary statistics from the full GWAS of Pasman et al. and bipolar disorder-summary statistics from the smaller GWAS of Stahl et al. To construct the genetic predictor, we identified SNPs that were both genome-wide significant (P<5e-8) for lifetime cannabis use and present in the bipolar study. Then, we excluded SNPs with minor allele frequency (MAF) < 0.01, sample size below 100,000 (lifetime cannabis use) or 40,000 (bipolar disorder), info score below 0.8, that were ambiguous (had alleles A&T or C&G) or were not consistent with the SNPs in 1000 Genome Project (21). Finally, we thinned so that no pair of SNPs remained within 3 centimorgan with correlation-squared r^2^ > 0.05.

Next, we investigated whether bipolar disorder is a causal risk factor for cannabis use. Likewise, we identified SNPs that were genome-wide significant (P<5e-8) for bipolar disorder, for which we also had summary statistics for cannabis use. For this we used summary statistics from the full GWAS of Stahl et al. (for bipolar disorder) and summary statistics from the smaller GWAS of Pasman et al. (for lifetime cannabis use). We did not filter the bipolar summary statistics based on sample size as these were not reported.

To perform the MR analyses (i.e. test whether the genetic predictor for lifetime cannabis use was associated with bipolar disorder, then whether the genetic predictor for bipolar disorder was associated with lifetime cannabis use), we used the R package “Mendelian-Randomization” (22). For the main analysis, we used inverse-variance regression to estimate the regression slope (a significantly non-zero slope indicates that the exposure is causal for the outcome). We also performed two sensitivity analyses, weighted-median regression (which gives unbiased estimates of the slope provided at least half the SNPs are valid instrumental variables) and Egger regression (which tests for the presence of pleiotropy).

## Results

We identified six independent, genome-wide significant SNPs for lifetime cannabis use, which in total explained 0.17% of phenotypic variation. As reported in Figure 1 and Table 1, we did not find evidence for a causal influence of liability to cannabis use on bipolar disorder. We identified 17 independent, genome-wide significant SNPs for bipolar disorder, which in total explained 0.29% of phenotypic variation. As reported in Figure 2 and Table 1, inverse-variance regression concludes that there is a causal effect of liability to bipolar disorder on lifetime cannabis use. The conclusion was the same if we instead used weighted-median regression, while Egger regression did not find significant evidence for pleiotropy.

**Table 1.** The causal relationship between lifetime cannabis use and bipolar disorder. Results from the two methods: inverse-weighted regression (IVW) and weighted median regression. The estimates of the slope (SD) and p-value are reported in column 3-4. Significant tests (P<0.05) are marked in bold.

| Exposure → Outcome | Method | Slope (SD) | P-value |
| --- | --- | --- | --- |
| Lifetime cannabis use → Bipolar disorder | IVW | 0.168 (0.180) | .351 |
|  | Weighted median | 0.249 (0.148) | .093 |
| Bipolar disorder → Lifetime cannabis use | IVW | 0.174 (0.039) | **.00001** |
|  | Weighted median | 0.168 (0.052) | **.001** |

**Figure 1:**
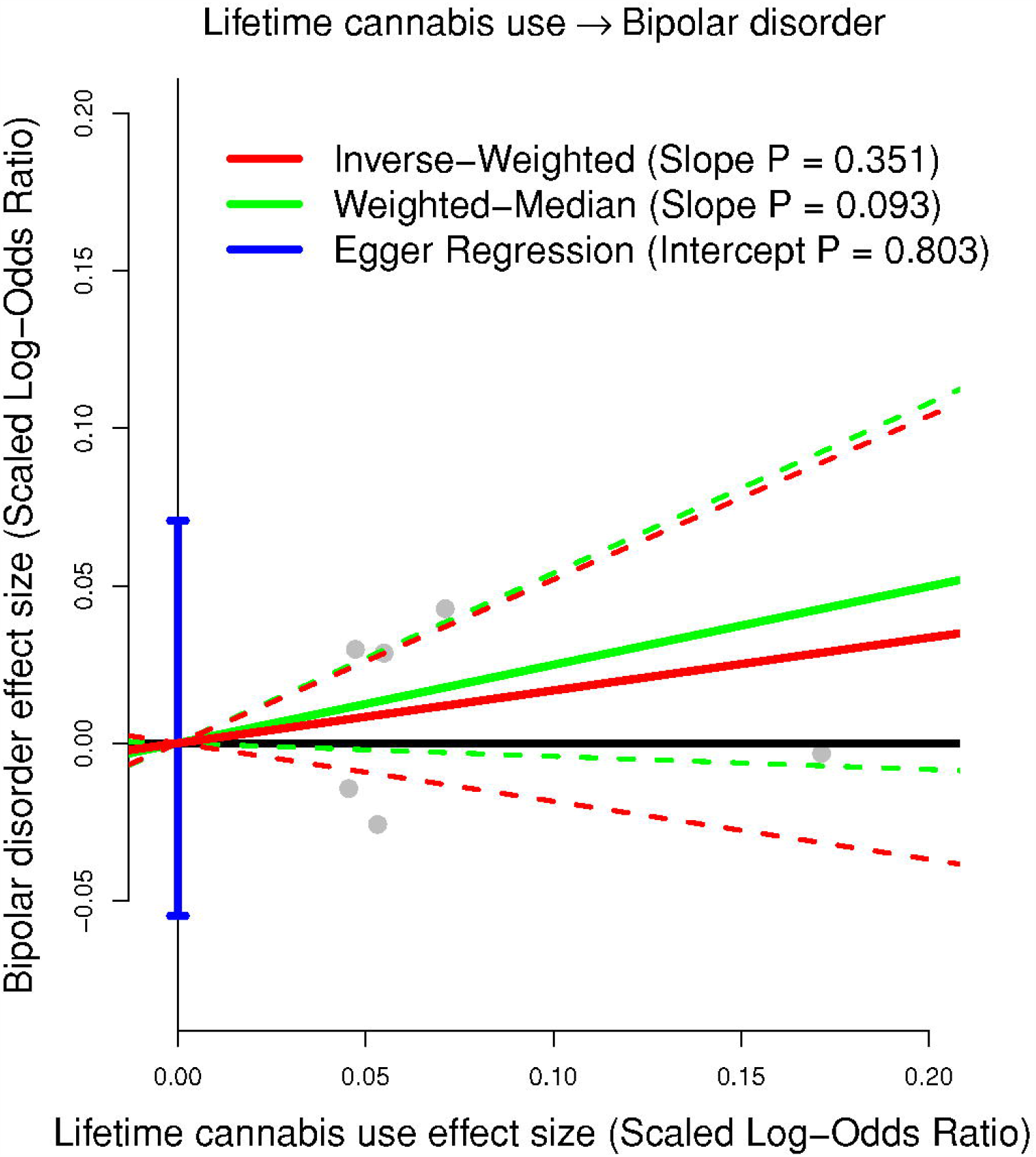
The figure plots the lifetime cannabis use effect sizes (scaled log-odds ratio) on the x-axis against the bipolar disorder effect sizes (scaled log-odds ratio) on the y-axis. The slope (and corresponding 95% confidence intervals) through the 6 independent, genome-wide significant SNPs is estimated using inverse-variance regression (red solid and dashed lines), weighted-median regression (green solid and dashed lines) and Egger regression (blue solid and dashed lines). The blue vertical and black horizontal lines indicate the 95%-confidence interval for the intercept from Egger regression and no effect, respectively.

**Figure 2:**
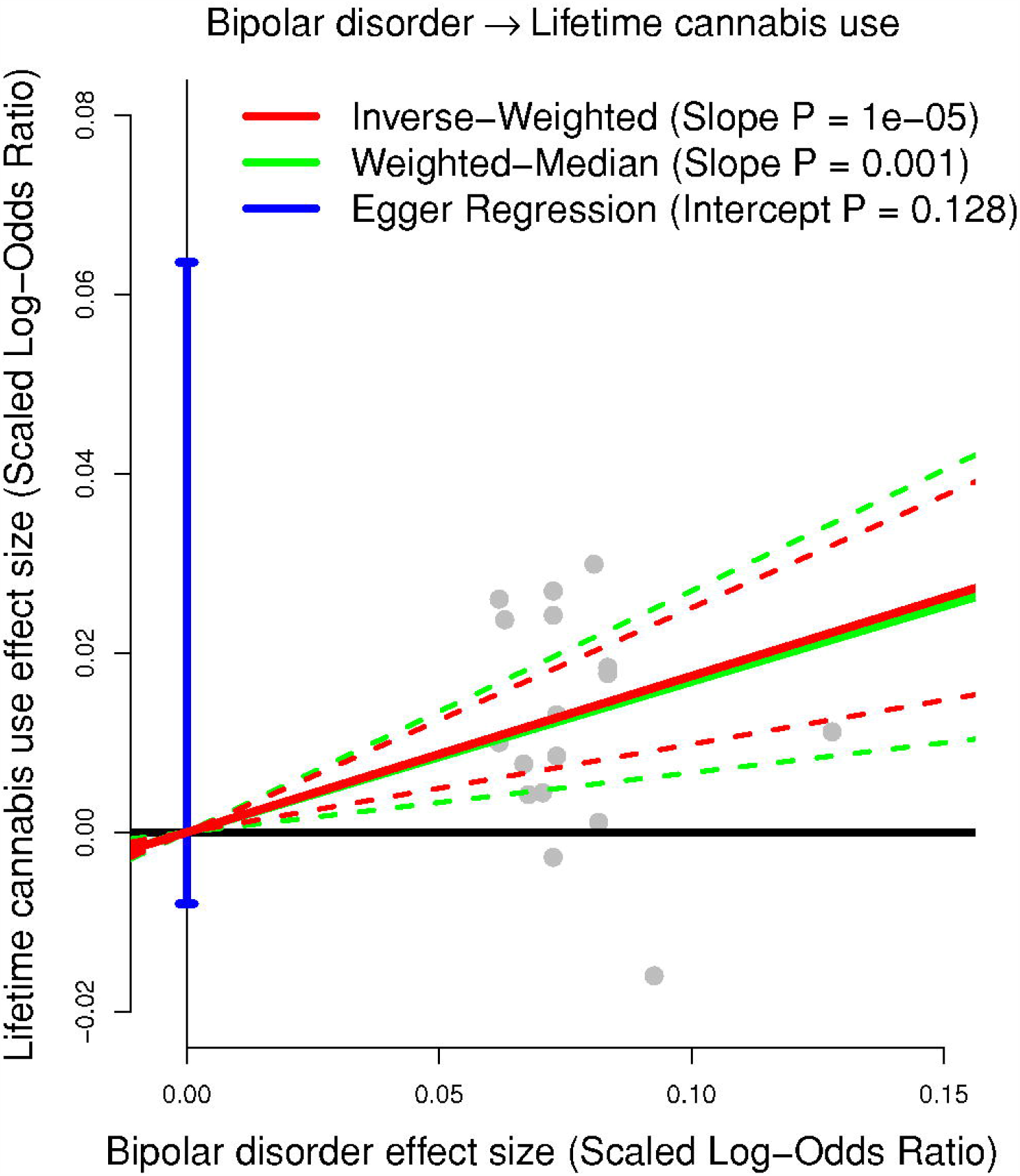
The figure plots the bipolar disorder effect sizes (scaled log-odds ratio) on the x-axis against the lifetime cannabis use effect sizes (scaled log-odds ratio) on the y-axis. The slope (and corresponding 95% confidence intervals) through the 6 independent, genome-wide significant SNPs is estimated using inverse-variance regression (red solid and dashed lines), weighted-median regression (green solid and dashed lines) and Egger regression (blue solid and dashed lines). The blue vertical and black horizontal lines indicate the 95%-confidence interval for the intercept from Egger regression and no effect, respectively.

## Discussion

In the present study we used MR to investigate the relationship between bipolar disorder and lifetime cannabis use. We found evidence for a causal effect of bipolar disorder on lifetime cannabis use, but not vice versa. These findings indicate that individuals with a higher genetic liability to bipolar disorder have a higher risk of cannabis use, which explain part of the association observed between bipolar disorder and cannabis use.

No previous MR study has investigated the relationship between cannabis use and bipolar disorder. However, our results are partly consistent with MR studies on cannabis use and other mental disorders. Specifically, these studies have reported that the liability to schizophrenia (9), depression (12), and ADHD (13), respectively, all increase the risk of cannabis use, but found no significant effects in the other direction (cannabis use → psychiatric disorder). In contrast, both Vaucher *et al*. and Gage *et al*. found that use of cannabis was associated with an increased risk of schizophrenia (10, 11).

The present findings suggest that liability to bipolar disorder increases the risk of using cannabis at least once. This causal effect may have several possible explanations. First, the causality may be mediated by (prodromal) manic symptoms; states of impulsivity, sensation-seeking, or risk taking behavior may all increase the risk of exposure to environments with more cannabis, and /or increase the risk of trying cannabis in any environment (23, 24). Second, the genetic liability to bipolar disorder is partially overlapping with the genetic liability to certain personality traits such as openness, and extraversion (25), both of which are also genetically overlapping with lifetime cannabis use (9). Therefore, the increased risk of lifetime cannabis use may be mediated by (non-pathological) personality traits associated with bipolar disorder liability and not bipolar disorder itself. Dissecting genetic liability to bipolar disorder from liability to personality traits may be of interest for future studies but is outside the scope of the present study. Another hypothesis that is often brought up in the context of substance abuse and psychiatric disorders is the self-medication hypothesis: that the experience of symptom relief from using cannabis causes patients to use more cannabis. However, this hypothesis cannot explain the present findings as the effect (lifetime cannabis use, yes /no) would thus precede the cause (experiencing symptom relief), representing an invalid causal model. Self-medication may therefore explain why some patients *continue* to use cannabis, but not why they try using cannabis in the first place, unless the recall of having ever used cannabis is positively enforced by having used a larger amount of cannabis (as self-medication), or if patients are advised to try cannabis as self-medication by others.

The present study has some limitations. The use of genetic instruments to test causal effects for an exposure on an outcome, requires that the following key assumptions are met: (a) the SNPs are predictive of the exposure variable, (b) the SNPs are not associated with a confounder of the exposure-outcome association, (c) the SNPs only affect the outcome through the exposure variable (i.e. there is no directional pleiotropy). We are confident of (a), because our genetic predictors only used SNPs robustly (P < 5e-8) associated with the exposures, and (b) because SNPs are randomly allocated during meiosis. It is impossible to directly test (c), however our two sensitivity analyses suggest that our findings are not the result of pleiotropy. Further, we observed that none of the 6 genome-wide significant SNPs for cannabis were causally associated with bipolar disorder (Bonferroni-corrected P < 0.05 /6), and likewise none of the 17 genome-wide SNPs for bipolar disorder were causally associated with lifetime cannabis use (Bonferroni-corrected P < 0.05 /17).

The lack of evidence for a causal effect of cannabis use on bipolar disorder risk may be due to insufficient power, as the genetic instruments for lifetime cannabis use were weak (only explaining ∼17% of the variance) and the investigated effect size may be small. Several observational studies suggest that while *heavy* cannabis use may carry a significant risk of developing bipolar disorder, the effect of having ever (vs. never) used cannabis may be negligible (26, 27). This dose-dependency for cannabis and bipolar disorder has also consistently been shown for cannabis and psychosis (3). Consequently, the present results neither support nor deny a causal effect of cannabis use on the risk of bipolar disorder. Furthermore, the present study cannot provide evidence regarding the effect of bipolar disorder liability on the risk of cannabis *dependence*, as the link between cannabis use and cannabis dependence is uncertain. The general probability for cannabis users to transition into cannabis dependence has been estimated to be 8-9% 10 years after first use (28). This transition-risk could however be enhanced in patients with mood disorders (29-31), a highly clinically relevant possibility, which should be subjected to further study.

In conclusion, our findings indicate that the liability to bipolar disorder increases the risk of lifetime cannabis use (trying cannabis at least once). This effect explains at least a part of the association between cannabis use and bipolar disorder. An effect in the other direction – i.e. whether liability to cannabis use increases the risk of bipolar disorder – can neither be confirmed nor refuted based on the present results. This new knowledge represents an important addition to the understanding of the complex relationship between cannabis use and bipolar disorder.

## Data Availability

The present study did not generate original data, but analyzed publicly available data, as referenced in the manuscript.

## Acknowledgments

This study included summary statistics of a genetic study on cannabis use (Pasman et al, in press Nature Neuroscience). We would like to acknowledge all participating groups of the International Cannabis Consortium, and in particular the members of the working group including Joelle Pasman, Karin Verweij, Nathan Gillespie, Eske Derks, and Jacqueline Vink. Pasman et al, (2018) included data from the UK Biobank resource under application numbers 9905, 16406 and 25331.

## Competing interests

The authors declare no competing interests

## Notes

### Competing Interest Statement

The authors have declared no competing interest.

### Funding Statement

No funding was received

